# The benefits and harms of oral iron supplementation in non-anaemic pregnant women: A systematic review and meta-analysis

**DOI:** 10.1101/2024.02.13.24302756

**Authors:** Archie Watt, Holden Eaton, Kate Eastwick-Jones, Elizabeth Thomas, Annette Plüddemann

## Abstract

**Objective:** Iron deficiency during pregnancy poses a significant risk to both maternal and foetal health. Despite increased iron requirements during pregnancy, current UK NICE guidelines do not give clear advice on antenatal iron supplementation for non-anaemic women. We aimed to assess whether the benefits of routine antenatal supplementation outweigh potential harms for non-anaemic women.

**Methods:** The Cochrane Library, MEDLINE, Embase and clinical trial registries were searched for randomised control trials (RCTs) and observational studies comparing oral iron supplementation with placebo or no supplement in non-anaemic pregnant women. The relevant data were extracted, and the risk of bias for included studies was assessed using the Cochrane Risk of Bias tool and the Newcastle-Ottawa Scale. Where appropriate, meta-analysis was conducted using ‘R’.

**Results:** 23 eligible studies were identified including 4492 non-anaemic women who were followed through pregnancy. Haemoglobin and ferritin levels were consistently higher in individuals receiving iron compared with control groups, although both findings were associated with a high degree of heterogeneity (I^2^ = 92% and 87% respectively) and therefore did not warrant a pooled analysis. Iron supplementation was associated with a significant reduction in rate of maternal anaemia (OR = 0.36; 95% CI = 0.22 - 0.61, p<.001; I^2^ = 54%; moderate certainty, NNT 8). There was no significant effect of intervention on birth weight (MD = 22.97g, 95% CI = -56.27 to 102.22, p = 0.57; I^2^ = 64%; very low certainty). Of the 18 studies reporting adverse effects, none found a significant influence of supplementation on GI disturbance, caesarean sections or preterm births.

**Conclusions:** Prophylactic iron supplementation reduces the risk of maternal anaemia in pregnancy. Limited evidence was found relating to the harms of supplementation in non-anaemic pregnant women, highlighting the need for further research to inform practice guidelines and support clinical decision making.

**Registration:** The study protocol was registered on the Open Science Framework (DOI 10.17605/OSF.IO/HKZ4C).

**Key Points:** **What is this research focused on exploring, validating, or solving?**

Antenatal iron supplementation is known to benefit pregnant women with iron deficiency anaemia, resulting in improved maternal and foetal outcomes. We explored whether these beneficial effects extend to non-anaemic pregnant women and whether they outweigh potential harms of supplementation.

**What conclusions did this research draw through design, method, and analysis?**

We have shown that supplementation of non-anaemic women helps prevent maternal anaemia and increases maternal haemoglobin. We have also identified a significant paucity in available evidence surrounding side effects of iron supplementation.

**What is the value, meaning and impact of your research? Is there any followup study based on this research?**

By clarifying the benefits of supplementation, we hope to assist decision making in primary care. This is particularly relevant given the current discrepancies in international guidelines. Our findings strengthen the evidence base in favour of universal supplementation, but focused research into side effects is still required to better qualify risk.

## Introduction

The UK prevalence of iron deficiency anaemia (IDA) in pregnant women is 23%^1^. IDA can negatively impact both maternal health and foetal health and development, increasing risk of premature birth, postpartum depression and low newborn birth weight^2^. There is also increasing evidence of maternal and foetal harms even before iron deficiency (ID) progresses to IDA^3^. Infants born of ID mothers may develop impaired cognitive, motor and social-emotional function^4,5^. As many as 91% of women experience ID during pregnancy^6^.

Current guidance advises that pregnant women with IDA or ID should take iron supplements^7^. However, iron parameters (e.g. serum ferritin) are not routinely assessed in pregnancy, unless the mother is deemed clinically ‘at-risk’ of anaemia^7^. This lack of testing means pregnant women with inadequate iron stores may not be identified. In addition, women who are initially iron replete can develop ID or IDA as they progress through pregnancy. These groups of women, who are not currently recommended iron supplements in the UK, may benefit from additional iron intake (figure 1).

**Figure 1:**
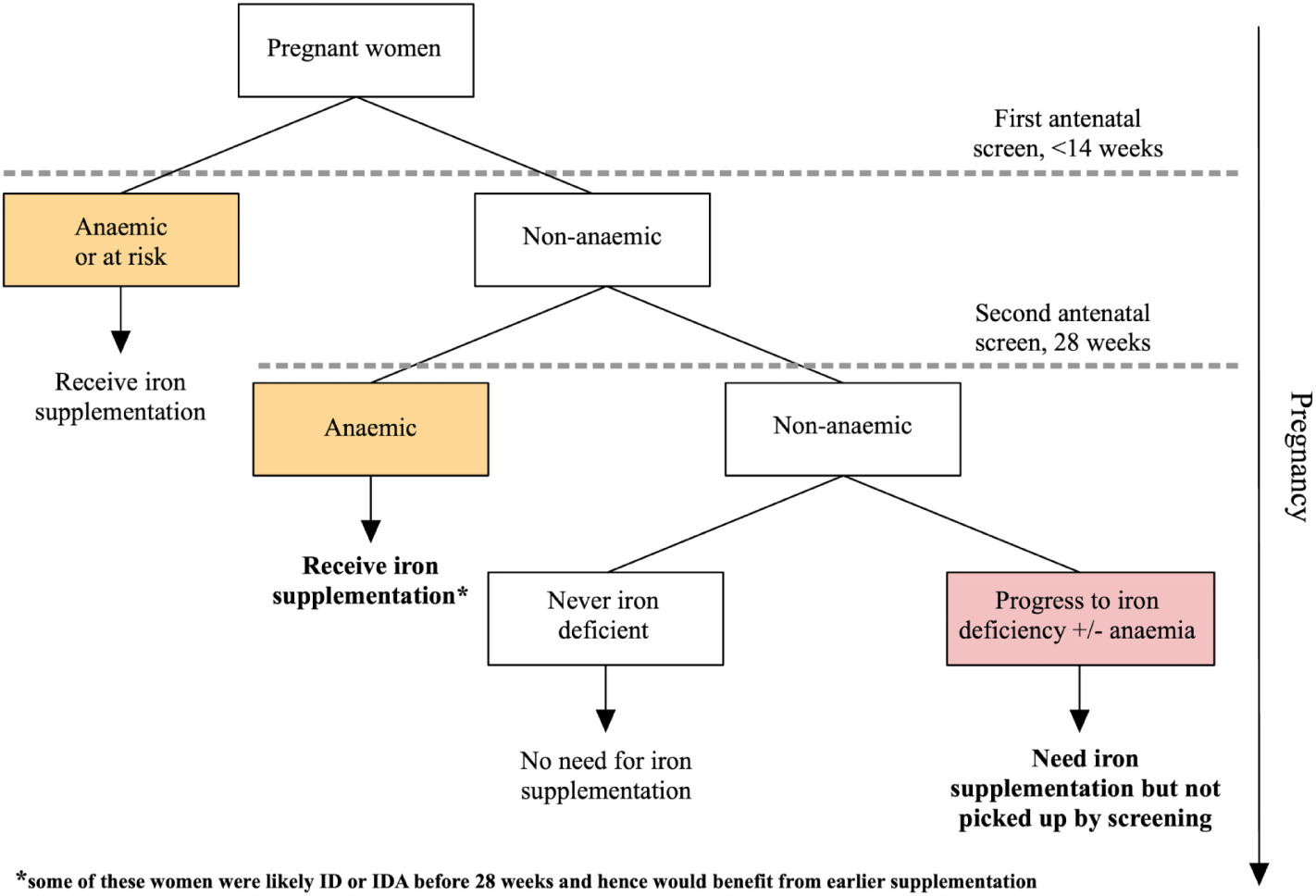
infographic detailing iron and Hb status through pregnancy as per current practice and guidelines.

The World Health Organisation and other international guidelines advise that all pregnant women, regardless of iron status, receive iron supplementation alongside folic acid^8–11^. However, UK guidelines remain ambiguous. The Royal College of Obstetricians and Gynaecologists recommends against universal supplementation^12^. Likewise, until April 2023, the National Institute for Health and Care Excellence (NICE) recommended against prophylactic iron supplementation in women without established IDA. Updated guidance advises that women already taking prophylactic iron supplements should continue to do so during pregnancy^1^, but still fails to address the rest of the pregnant population.

Arguments against routine prophylactic iron supplementation in pregnancy suggest that the harms outweigh the benefits in non-anaemic women. For example, iron overload and high haemoglobin may be associated with adverse outcomes including preterm birth and foetal growth restriction^2,13,14^. In addition, gastrointestinal side effects are commonly reported by women. However, the harm of iron supplementation to non-anaemic pregnant women has not been clearly quantified in the literature. Given the growing body of evidence that ID and IDA cause maternal and foetal harm, it is critical to reexamine whether iron supplementation provides a net benefit to this population’s health and wellbeing.

A 2015 review concluded that supplementation reduced maternal anaemia at term by 70%, and improved foetal outcomes, such as birth weight^15^. However, this review explored the effect for all pregnant women rather than non-anaemic women specifically. Since women with IDA at the start of pregnancy are already offered iron supplementation in the UK, the findings cannot be used to determine the benefit of universal, rather than selective, iron supplementation. A 2023 systematic review examined only non-anaemic women, but specified that participants must also be iron-replete^16^. Again, the findings in this population are less relevant to the UK context since iron status is not routinely investigated. Our review will explore both the benefits and harms of iron supplementation in non-anaemic pregnant women, a key population in the discussion surrounding routine iron supplementation in the UK.

## Methods

The study protocol was registered on the Open Science Framework (DOI 10.17605/OSF.IO/HKZ4C). This review is reported according to PRISMA (preferred reporting items for systematic reviews and meta-analyses) reporting standards ^17^.

### Patient and Public Involvement

Patient encounters in primary care inspired initial investigation and research into this topic. However, patients and the public were not involved in the undertaking of this study.

### Data sources and searches

The Cochrane Library, MEDLINE and Embase (via Ovid) were searched for randomised clinical trials (RCTs) and observational studies published up to 31/01/2024 including non-anaemic pregnant women comparing oral iron supplementation to no supplementation or placebo. Both clinicaltrials.gov and who.int/clinical-trials-registry-platform were searched for recently completed and ongoing studies. Details of search terms used are described in supplement 1 (S1). The reference lists of included papers and recent systematic reviews were also screened.

### Selection criteria

RCTs and observational studies were included that compared oral iron supplementation with no supplementation, placebo or supplementation without iron in non-anaemic (according to pre-supplementation Hb measurement), iron-replete pregnant women.

Studies of oral iron tablets containing only iron or iron as part of a multivitamin were included. Studies were excluded if the supplementation was a dietary change alone or not administered orally. Studies were not excluded based on iron dosage or formulation. Studies involving co-interventions were included only if both groups received the same co-intervention. No restriction was placed on publication year or language.

### Outcomes

### Primary outcomes

- Maternal: Haemoglobin and serum ferritin at term or postpartum.
- Infant: Birthweight.
- Reporting of harms/side effects.

### Secondary outcomes

- Maternal: Quality of life, fatigue, gastrointestinal side effects, gestational diabetes, infections in pregnancy, other side effects.
- Infant: Birth complications, preterm birth.

### Study selection

Two reviewers independently screened titles, abstracts and full texts of the search results using the Rayyan QCRI online software^18^. Records from clinical trial registries were screened by one reviewer and any disagreements were discussed among the three reviewers until a consensus was reached.

### Data extraction

If a study outcome was reported in more than one publication, data was only extracted from the publication with the most comprehensive data. Data was extracted relating to study design, location, number of participants and their population characteristics, baseline and post-intervention Hb and iron status, treatment and dose. When needed, authors were contacted for any additional data that was required.

### Assessment of risk of bias and quality of evidence

Included studies were independently assessed by two reviewers for risk of bias using the Cochrane risk of bias tool for RCTs^19^ (RoB2), or the Newcastle Ottawa Scale for observational studies^20^ (NOS). Any disagreement was resolved by discussion.

The GRADE (Grades of Recommendation, Assessment, Development and Evaluation) approach was used to assess the quality of the evidence^21^.

### Data analysis

#### Measures of treatment effect

For dichotomous data, results were presented as summary odds ratio (OR) with 95% confidence intervals (CI). For continuous data, the mean difference (MD) was used.

#### Unit of analysis issues

Cluster-randomised trials or cross-over trials were not included. Where studies measured outcomes at multiple time points, only the measure recorded closest to 40 weeks of gestation was used for analysis. Where studies had multiple intervention or control groups, they were combined using the method in Cochrane’s handbook^22^.

#### Assessment of reporting biases

Publication bias was investigated on outcomes with more than 10 trials by examining the funnel plots for asymmetry. Alternative reasons were considered for asymmetry, including differences in study methodological quality and true heterogeneity in intervention effects.

#### Data synthesis

Statistical analysis was carried out using the meta package in R version 4.0.5 ^23^.

Trial results were pooled using a random-effects model since there was a high degree of methodological heterogeneity. The random-effects model gives the average intervention effect assuming a distribution of different but related intervention effects for each trial.

For incidence of anaemia data synthesis, Peto’s method was used since several studies found zero events in one or both groups.

#### Assessment and investigation of heterogeneity

Heterogeneity was assessed and quantified using Chi^2^, I^2^ and Tau^2^ statistics in each analysis. We then investigated methodological and clinical sources of heterogeneity by conducting subgroup analysis based on the following variables:

1. Gestational age at start of supplementation: early (<20 weeks gestation), late (>20 weeks gestation) or mixed.
2. Dose of iron: Low (<30mg), Medium (30-60mg) or High (>60mg)
3. Definition of non-anaemic: below WHO (cut-off below 110g/L), WHO (cut-off 110g/L) or above WHO (cut-off above 110g/L).
4. Timing of outcome measurement during pregnancy: before 34 weeks, after 34 weeks or at term.

Where there was evidence of a difference between subgroups in analyses with at least 10 studies, we presented the Chi^2^ statistic and P value, and the interaction I^2^ value.

#### Sensitivity analysis

Sensitivity analysis was conducted by removing studies determined to be high risk of bias.

#### Post hoc changes to the protocol

Several studies measured Hb during pregnancy and treated anaemic participants. Whether the participant was excluded or included, this would reduce the apparent effect of supplementation. To overcome this limitation, we reported the incidence of anaemia as an additional post-hoc outcome. Incidence of anaemia is also more clinically relevant than Hb alone.

Birth weight was used instead of ‘size for gestational age’ as a measure of foetal outcomes, as this was much more commonly reported.

## Results

### Study selection

3,467 articles were identified by the search following de-duplication. 26 eligible articles were identified for inclusion (figure 2). Four more articles were identified from reference lists of included studies and relevant systematic reviews. Therefore, in total, 30 eligible articles were identified, corresponding to 23 unique studies - 21 RCTs and 2 observational studies.

**Figure 2:**
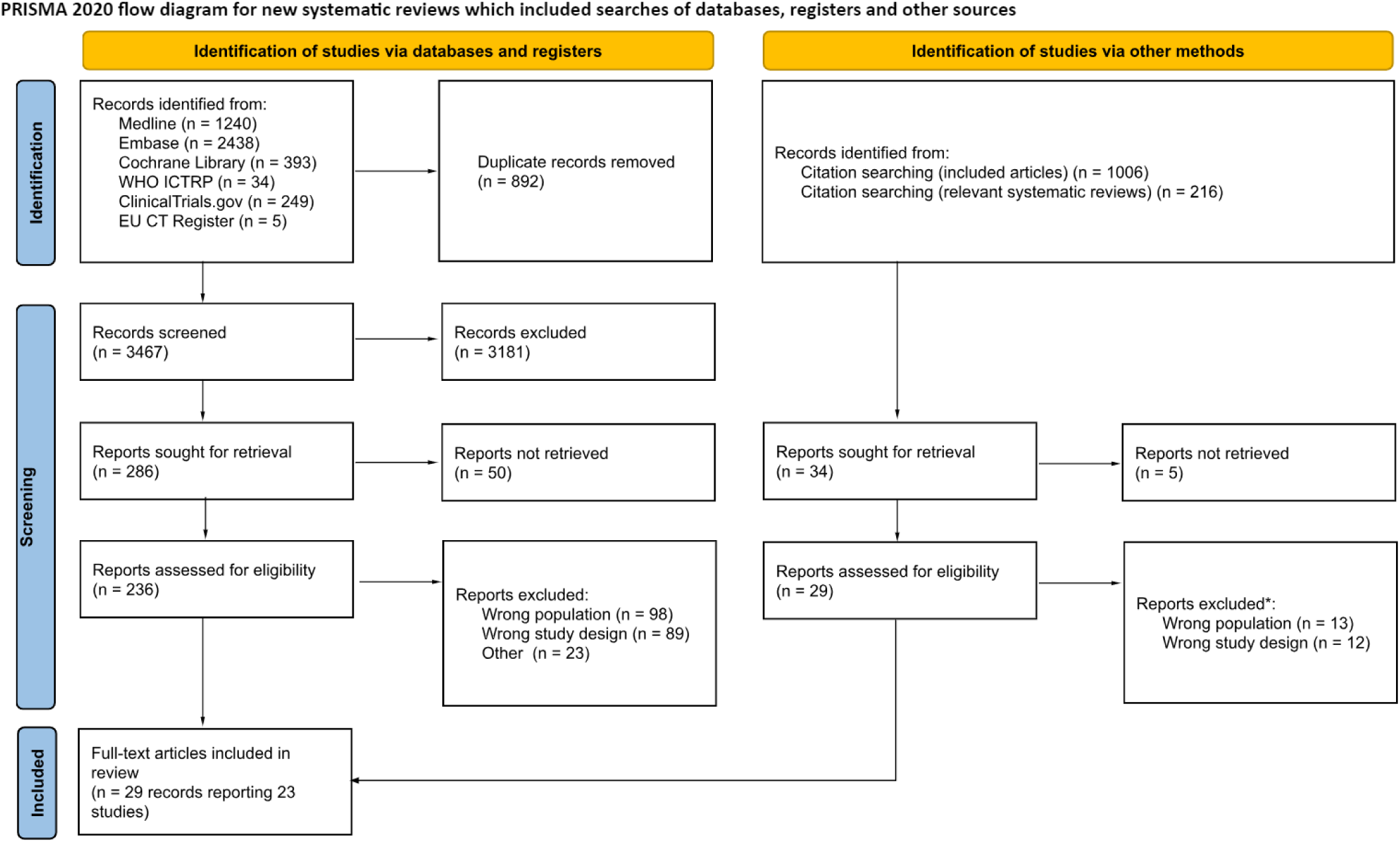
PRISMA study flow diagram ^17^.

### Study Characteristics

Out of 5678 women included at baseline across 23 studies, 4492 were followed throughout. Of these, 2296 received iron supplementation, 2050 received placebo and 155 were non-supplemented controls.

Population characteristics are shown in table S2. Three studies^24–26^ provided their own cut-off for non-anaemic patients which fell below the NICE definition, and 11 studies included a ferritin cut-off in their inclusion criteria^24,27–36^. These are reported in table S2.

Iron intervention was predominantly given in the form of ferrous sulphate, with doses varying from 20mg to 200mg once-daily. All but one study^33^ initiated supplementation at 20 weeks gestation or earlier, and all but two studies^28,30^ continued supplementation to term or later

### Risk of bias assessment

A risk of bias assessment was conducted for 21 of the studies (all unique RCTs included - figure 3). Overall risk was deemed as “Low” for only 2 studies, “Some concerns” for 13 studies and “High risk” for 6 studies. Understandably, many studies treated participants if their Hb dropped too low during the trial. Several studies excluded this data and so introduced a high degree of bias in favour of the null hypothesis^24,34,35,37,38^. Other studies analysed these participants in their original groups introducing less bias but by giving control participants iron they may have reduced the apparent effect of intervention^27,28^. One study excluded participants based on birth outcomes^31^ introducing a high degree of bias since these may be influenced by development of IDA. The most common reason for “some concerns” was absence of study pre-registration.

**Figure 3.**
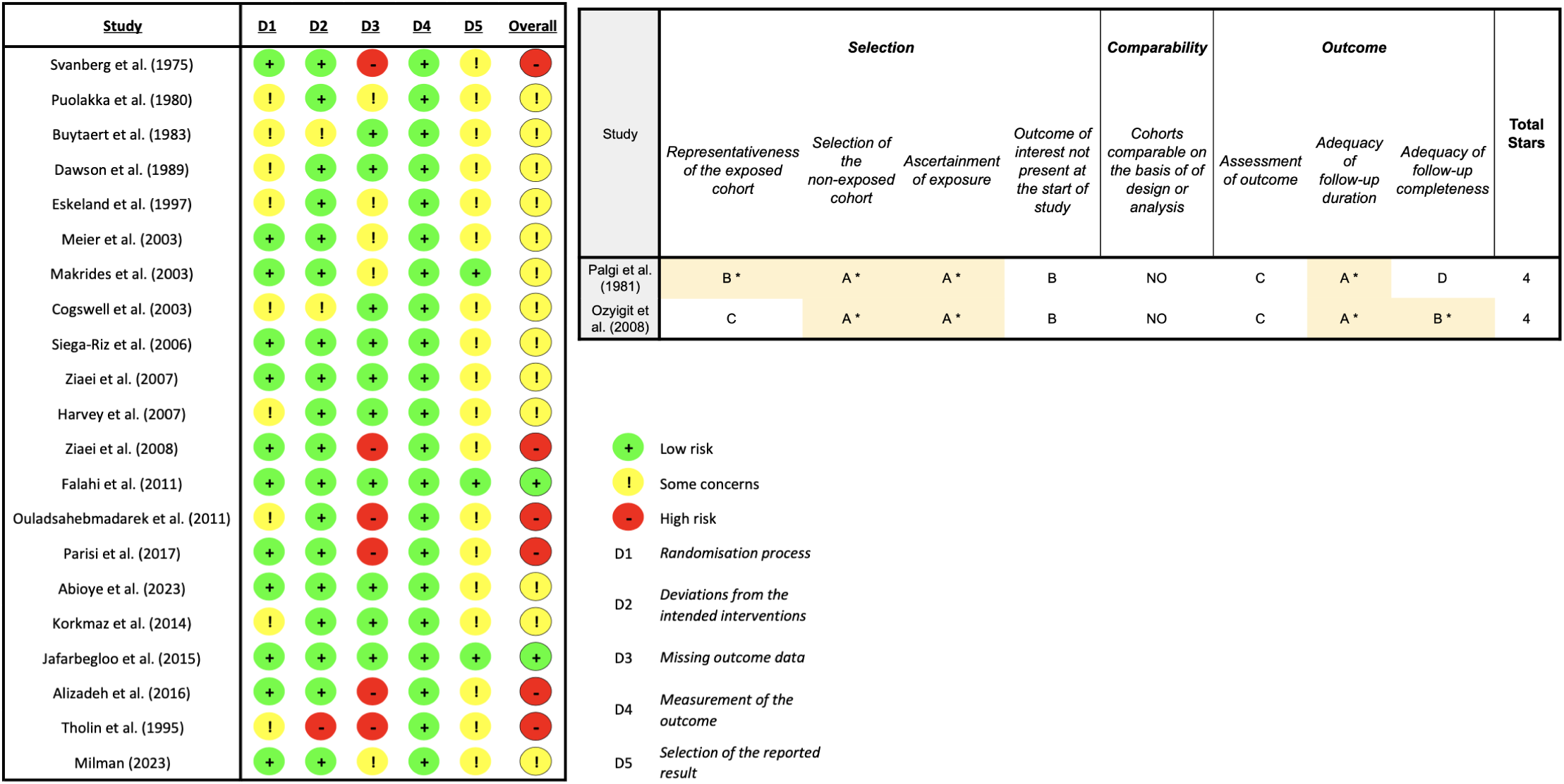
ROB2 assessment (left), Newcastle Ottawa scale (right)

Both included observational studies^26,39^ were assessed using the NOS tool (figure 3) and were scored as ‘fair quality’.

### Benefits of Supplementation

#### Haemoglobin

18 studies reported maternal Hb in the final trimester of pregnancy with a total of 4015 participants. 13 studies found that iron supplementation significantly increased maternal Hb compared to controls (Figure 4). The studies used a wide range of methodologies and since there was high heterogeneity in the results (I^2^ = 91%) we have not reported a pooled effect estimate. Effect sizes ranged from 0g/L to +17g/L. Subgroup analysis based on dose, the authors’ anaemia definition and supplement start time could not account for the heterogeneity (Figures S3, S4, S5). Studies measuring Hb before 34 weeks of pregnancy had significantly smaller effects of treatment than studies measuring at term or after 34 weeks (Chi^2^ = 14.55, p < .01) (Figure S6).

**Figure 4.**
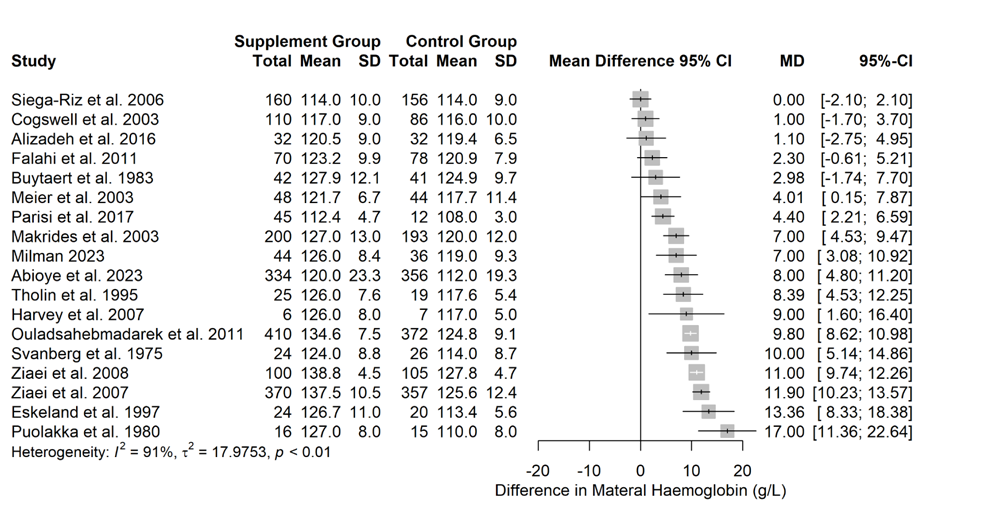
Forest plot showing the mean differences in maternal haemoglobin between iron supplemented and non-supplemented groups across 18 studies (n=4015). There was a high degree of heterogeneity across the studies (I^2^ = 91%) and so we have not reported a pooled estimate of effect. 13 of the studies reported statistically significant differences in haemoglobin and the effect size ranging from 0g/L to +17g/L.

Figure 4. - Effect of iron supplementation on maternal haemoglobin

#### Maternal Anaemia

We extracted incidence of maternal anaemia from 13 studies (n=2315). In the supplemented groups 128 of 1185 participants (10.8%) developed anaemia and in the control groups 231 of 1130 participants (20.4%) developed anaemia. The pooled odds ratio for developing anaemia was 0.33 (95% CI = 0.20 - 0.56, p<.001) (Figure 5). This means for every 1000 women given iron supplementation, 124 fewer cases of anaemia occurred. The certainty of evidence using GRADE assessment was moderate (downgraded for inconsistency and probable publication bias, upgraded due to the large effect).

**Figure 5.**
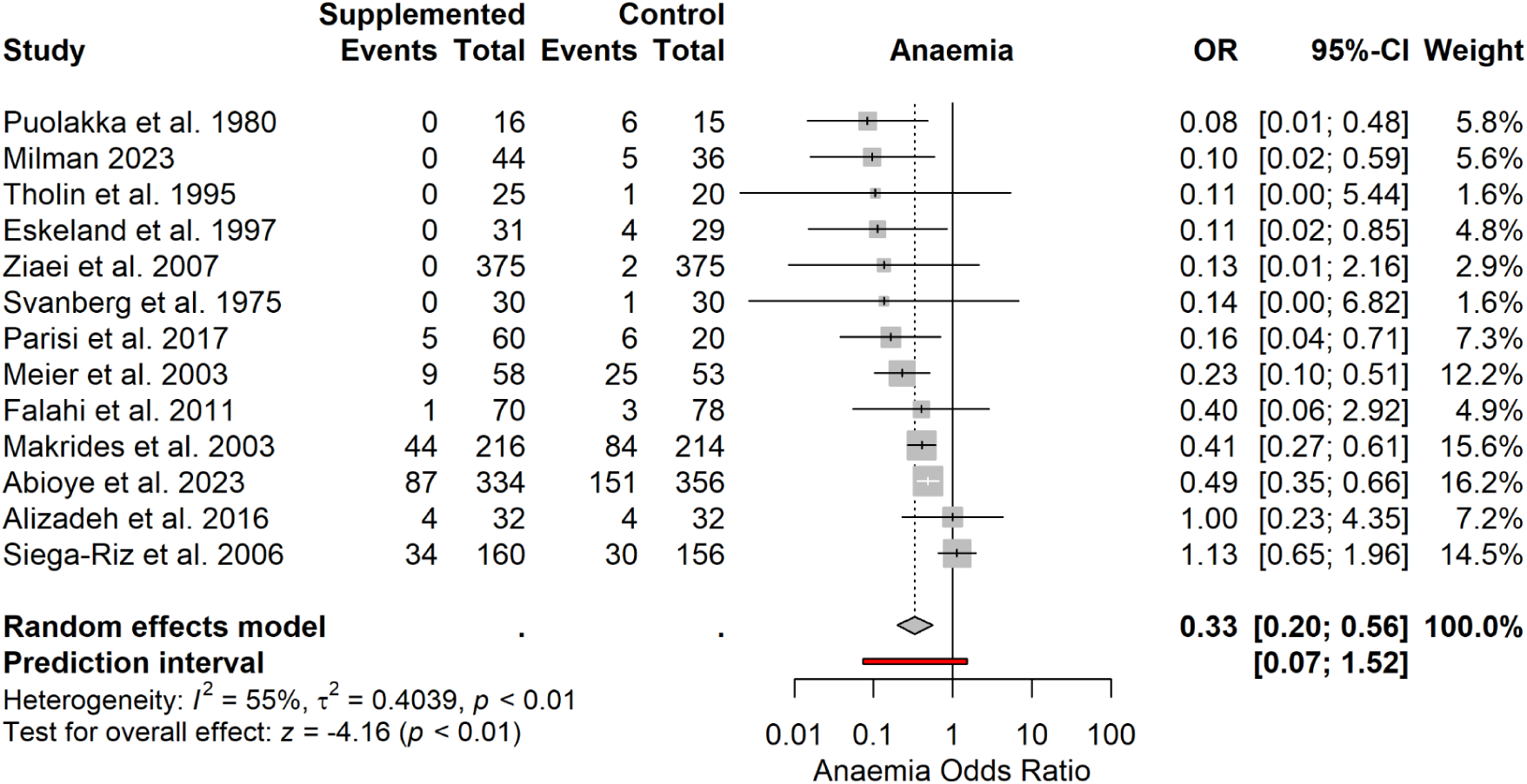
Forest plot showing the odds ratio of maternal anaemia between iron supplemented and non-supplemented groups across 13 studies (n=2315). There was a moderate degree of heterogeneity across the studies (I^2^ = 55%). The pooled odds ratio for developing anaemia was 0.33 (95% CI = 0.20 - 0.56, p<.001). This means for every 1000 women given iron supplementation, 124 fewer cases of anaemia occurred.

The results displayed moderate heterogeneity (I^2^ = 55%) although no studies reported anaemia risk was greater in the supplemented group. The heterogeneity was likely due to methodological differences although it was not explained through subgroup analyses based on dose, the authors’ anaemia definition and supplement start time (Figures S8, S9, S10). Subgroup analysis based on the time at which haemoglobin was measured eliminated heterogeneity for groups measuring late in pregnancy (I^2^ = 0%) and at term (I^2^ = 2%) (Figure S11). Publication bias was assessed via visual inspection of the funnel plot. There was apparent funnel plot asymmetry which could be due to publication bias or small study effects (Figure S12). Sensitivity analysis was conducted by removing studies with a high risk of bias. This did not affect the estimate of effect (OR = 0.33, 95% CI = 0.18 - 0.60, p<.001) (Figures S13).

Figure 5. - Effect of iron supplementation on risk of maternal anaemia

#### Maternal Ferritin

14 studies reported maternal ferritin with a total of 2853 participants. As with maternal haemoglobin there was a high degree of heterogeneity across the studies (I^2^ = 87%), although no studies found that supplementation reduced maternal ferritin (Figure S14). Due to the methodological heterogeneity we have not reported a pooled effect estimate. Effect sizes ranged from +1.28ng/mL to +42.00ng/mL.

#### Birth weight

13 studies reported birth weights for both intervention and control groups *(n=2994)*. Pooling the estimates showed no significant effect of intervention (MD = 17.75g, 95% CI = -55.74 to 91.24, p = 0.64) (Figure S15). There was a high degree of heterogeneity (I^2^ = 61%) and studies found both positive and negative effects of intervention. The certainty of evidence using GRADE assessment was very low (downgraded for very serious inconsistency and risk of bias).

### Harms of Supplementation

Of the 23 unique studies identified, 18 mentioned and reported at least one type of side effect.

#### GI side effects

Four papers reported on GI side effects^27,29,39,40^. These included nausea, vomiting, constipation, diarrhoea, abdominal pain, loss of appetite and heartburn. Palgi et al^39^ only reported side-effects in the intervention group, with no control group comparator. All three of the other studies reported no significant differences between intervention and control groups for any side effect.

#### Caesarean sections

Six studies reported C-section rates^27,29,31,41–43^. Of these, Makrides et al^29^ and Ziaei et al^43^ reported a non-significant difference between control and intervention groups. Eskeland et al^42^ did not report exact numbers, however reported a non-significant difference. The remaining studies did not conduct statistical comparisons although presented outcomes narratively. A random effects meta-analysis showed no significant effect of intervention (OR = 1.08, 95% CI = 0.84 to 1.38, p = 0.54 (Figure S16). The certainty of evidence using GRADE assessment was low (downgraded for imprecision and risk of bias).

#### Preterm births

Six papers reported on rates of preterm birth^28,30,32,33,38,43^. Pooled estimates showed no significant effect of intervention (OR = 0.81, 95% CI = 0.55 to 1.19, p = 0.28) (Figure S17). The certainty of evidence using GRADE assessment was moderate (downgraded for risk of bias).

#### Other side effects

Alongside the primary side-effects discussed above, papers also reported on a variety of other effects from iron supplementation. Due to their limited reporting, rather than meta-analyse these, we extracted the type of side effect reported to give an indication of what the literature has focused on to-date (a full list of these is included in Table 2). Three papers reported on aspects of zinc metabolism^25,29,41^, two on size of infant for gestational age^28,43^ (both finding significant effects in opposing directions), one on gestational diabetes^36^ and six on average gestational age at delivery^24,27,29,30,32,41^.

**Table 2.**
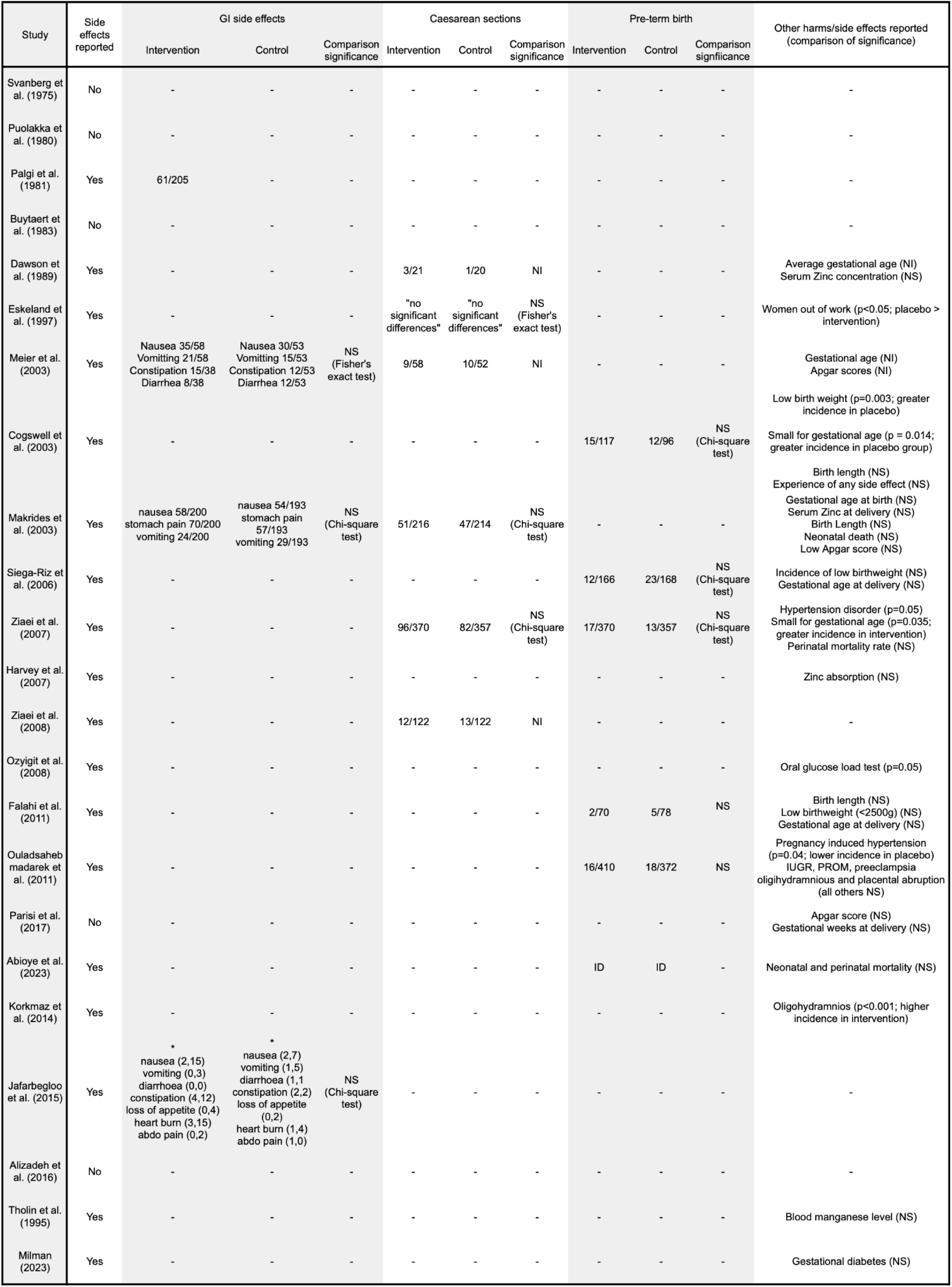
table of side-effects; ***\*\*\**** *= combined 3 means and SD using Repeating Cochrane’s formula;* ***NS*** *= non-significant result;* ***NI*** *= no information indicated;* ***ID*** *= insufficient data from paper to present figure;* ***\**** *= (x,y) indicates results at 24 and 32 weeks*

## Discussion

### Summary of findings

We have found that iron supplementation reduces the risk of anaemia in non-anaemic pregnant women. Supplementation improved haematological indices at term, including haemoglobin and serum ferritin. But we found no significant relationship between supplementation and foetal birth weight. Other maternal benefits including improved quality of life and reduced fatigue were not reported. Few included studies explored maternal harms. Across those that reported GI side effects, we found no significant association with iron supplementation. Foetal outcomes were better represented, with birth weight, preterm birth and average gestational age being explored in multiple studies. Our analysis did not highlight any significant difference in any foetal outcome.

### Findings in the context of current literature

A previous systematic review by Hansen et al found positive effects of daily iron on haemoglobin and serum ferritin at term in iron-replete, non-anaemic, pregnant women^16^. Similarly, Pena-Rosas et al found the same in pregnant women more generally^15^. Our results include data from non-anaemic women regardless of iron status. Given iron parameters are not routinely tested, this subpopulation is more relevant to the discussion surrounding universal prophylactic antenatal iron supplementation in the UK.

Gastrointestinal side effects are a commonly cited harm of oral iron supplementation^40^. These symptoms may arise due to incomplete absorption in the gut^44^ and could be sufficient to dissuade some pregnant women from initiating or continuing supplementation. A 2015 systematic review by Tolkien et al found a clear relationship between iron supplementation and GI-specific side effects in the non-pregnant population^45^. However, these findings may not translate to pregnant populations due to the increased iron requirement and altered physiology of pregnancy. In addition, single-armed studies may overestimate the prevalence of gastrointestinal side effects since these symptoms are common in pregnancy regardless of iron intake. We did not find a significantly increased rate of nausea, vomiting or diarrhoea in supplemented non-anaemic pregnant women. This seems to conflict with the anecdotal experiences of clinicians and pregnant women as well as the RCOG guidelines^12^. Our findings, based on the current evidence base, are inadequate to conclude whether such an association exists.

Other side effects of iron supplementation may result from downstream effects of iron overload^13^. Although it is unclear how oral iron overload might occur mechanistically, particularly in the context of tight homeostatic regulation of oral iron absorption, these micro-level effects are thought to translate to macro-systemic harms such as preeclampsia, prematurity, and foetal growth restriction^13,14^. Women with underlying genetic causes of excessive iron absorption, such as genetic haemochromatosis, may also be more at risk of iron-overload^11^, however no study we identified isolated these women specifically. We found no clear association between iron supplementation and preterm birth or birth weight following analysis. One study found a significant association between iron and pregnancy induced hypertension^38^ and another between iron and oligohydramnios^46^. Two studies found significant differences in proportions of births deemed ‘small for gestational age’ between groups, however the effects were in opposing directions^28,43^. Large variation in the type and detail of side effect reporting in included studies made it difficult to analyse whether such harms from supplementation are truly present. Finally, no study reported on the effect of iron supplementation on quality of life in the context of ID or IDA, despite its frequent mention in the literature^47^.

### Limitations of review

The included studies employed a variety of different methodologies, with differences in iron dose, formulation and regimen, timing of blood sampling, definition of anaemia and other variables. This heterogeneity limits the conclusions that can be drawn when trying to quantify the effect of iron supplementation. We could not identify a single variable which convincingly accounted for the heterogeneity in any outcome. Since the included studies were conducted in a variety of countries, we anticipate that differences in social and cultural context contributed to the heterogeneity. However, there was insufficient participant information to investigate this hypothesis.

Researchers had an ethical obligation to treat women in the control groups if they became anaemic before the end of the trial. These participants were either excluded or analysed in their original groups. Both of these options risk reducing the apparent benefit of iron supplementation compared to controls and may mean our findings underestimate the true effect of intervention. Some countries already recommend universal antenatal iron supplementation and so researchers can not conduct trials with unsupplemented controls. This limited the pool of available studies from which we could draw data.

### Implications for future policy and research

Despite identifying a clinically relevant benefit of iron supplementation for non-anaemic pregnant women, and highlighting the differences between UK and foreign guidelines^9–11^, it remains difficult to unequivocally recommend a change to NICE guidelines. Policymakers must take into consideration the needs of a specific population. For example, WHO guidelines must account for the large variation in baseline iron intakes across the globe^48,49^. A 2020 review by Milman et al investigating average maternal iron intake during pregnancy revealed variances even between European countries, both in terms of recommended daily intake and actual recorded intake^49^. This study indicated that the UK population most closely resembled those from Nordic countries, where iron supplementation is universally recommended.

Given the limitations in collecting data from countries with contrasting guidelines and differing populations, it is necessary to conduct large, randomised control trials within the UK, with a focus on quantifying side effects and subjective measures of maternal wellbeing. Observations from countries with similar population characteristics and iron intake, where iron supplementation is already routinely recommended, may also yield useful information to help inform UK-centric decisions.

## Conclusion

Non-anaemic pregnant women may use prophylactic iron supplementation to reduce their risk of anaemia. The exact magnitude of this effect will vary depending on individual context but we estimate that for every eight women taking iron supplementation, one person will be prevented from developing anaemia. We found little evidence regarding the harms of iron supplementation in non-anaemic pregnant women. It is not clear whether iron supplementation causes significant gastrointestinal disturbance, or any other proposed side effect, in this population. Researchers should consider investigating the tolerability and acceptability of iron supplementation to support clinical decision making. Routine iron supplementation during pregnancy may be appropriate, provided the risk of side effects and other harms can be more clearly quantified. This change to guidelines would bring the UK in line with other comparable countries.

## Author contributions

AW, HE and KEJ contributed equally to the study. AW, HE and KEJ developed the search terms and strategy with guidance from the institution’s academic librarian, carried out the searches, screening, extraction, analysis and interpretation of results in consultation with ET and AP. The final manuscript was written by AW, HE and KEJ, and reviewed by ET and AP.

## Data Availability

All data produced in the present work are contained in the manuscript

https://osf.io/hkz4c/

## Supplementary Materials

**Figure S1.**
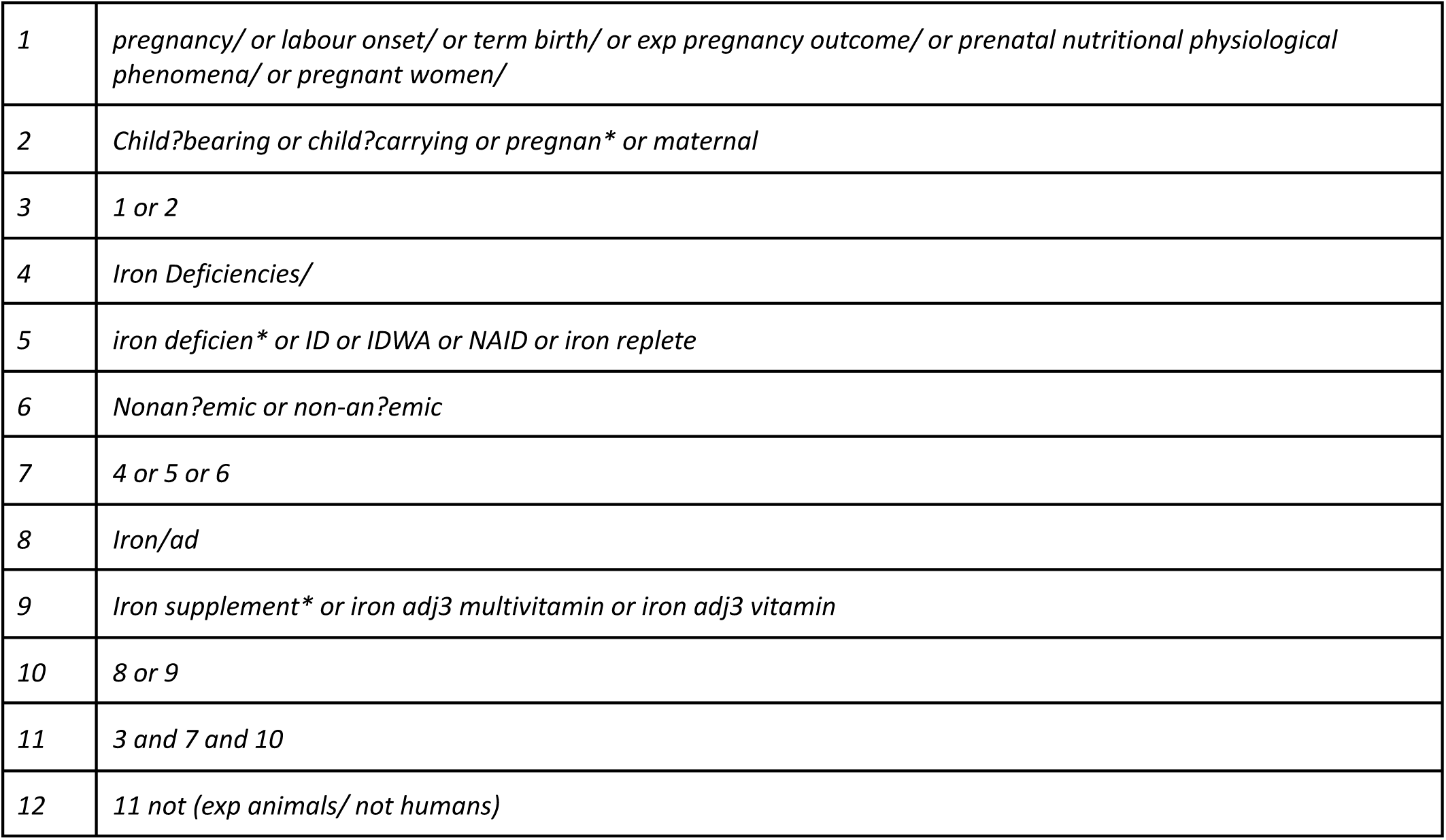
Table of search strategy.

**Table S2.**
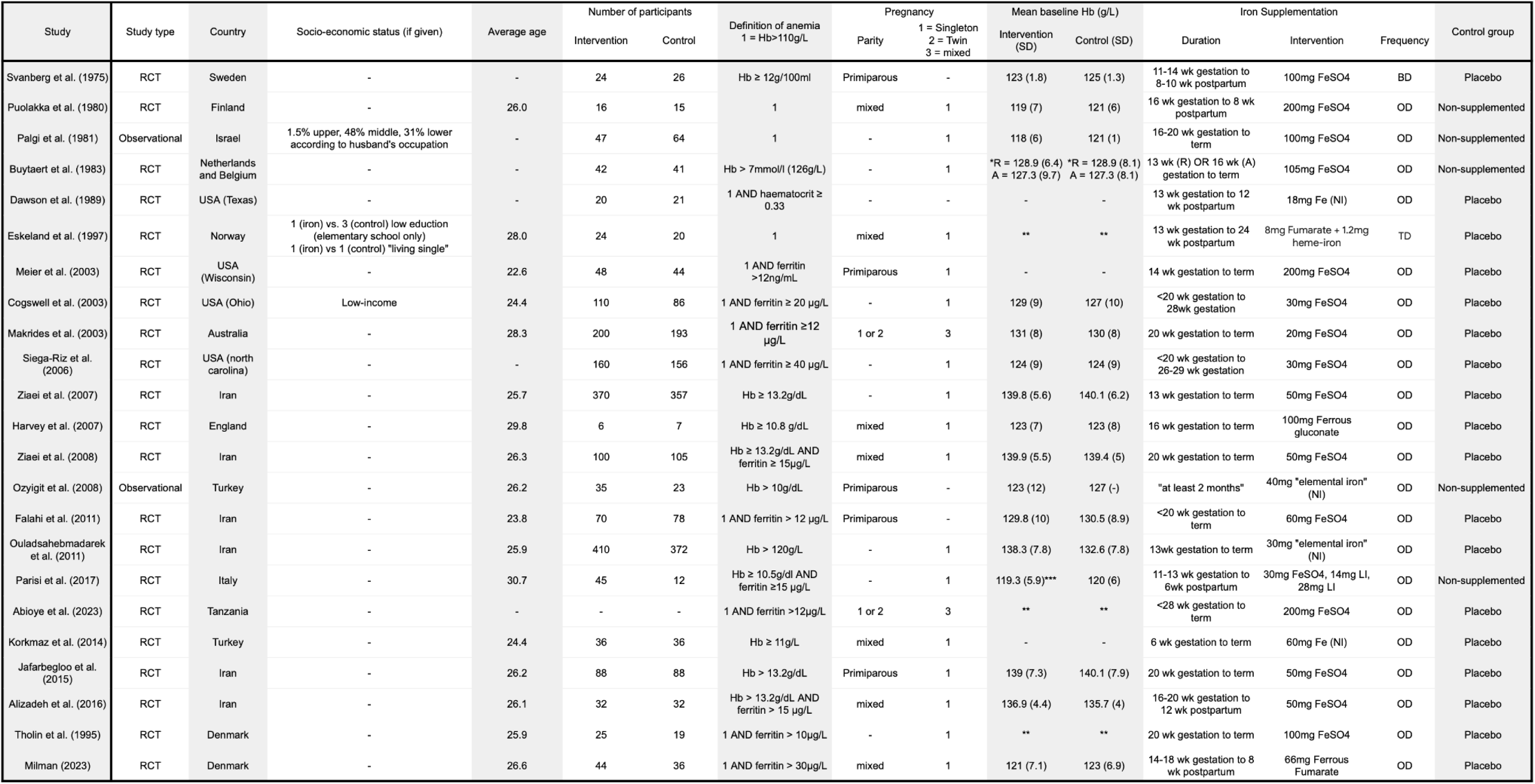
table of study characteristics; **-** indicates no information provided; **\*** study conducted with two separate groups, A = Antwerp, R = Rotterdam; **\*\*** Only median value available; **NI** iron type not specifically indicated; **LI** liposomal iron (Sideral R.M. Pharmanutra, Pisa PI, Italy ***\*\*\**** *Combined 3 means and SD using Repeating Cochrane’s formula* ***OD, BD, TD*** *Once Daily, Twice Daily, Thrice Daily*

**Figure S3.**
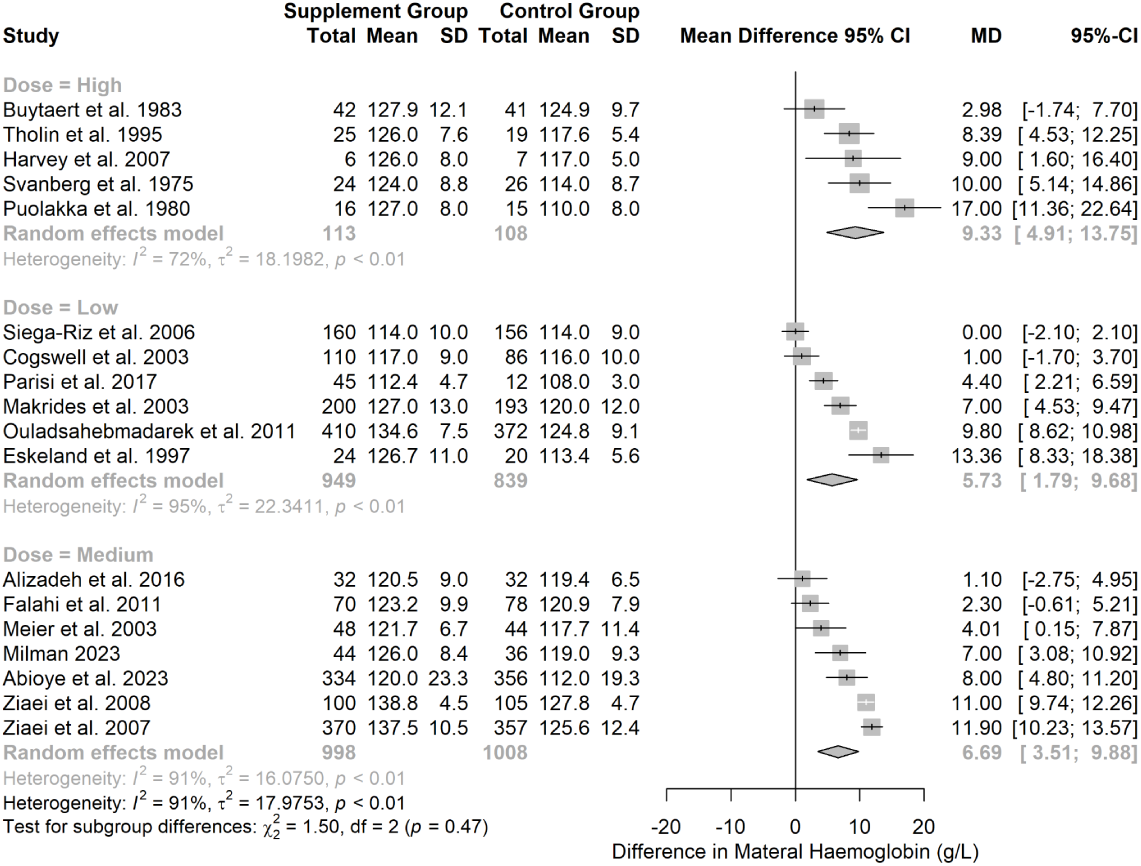
Forest plot showing effect of iron supplementation on maternal haemoglobin with subgroup analysis based on supplement dose (<31mg, 31mg to 60mg, >60mg).

**Figure S4.**
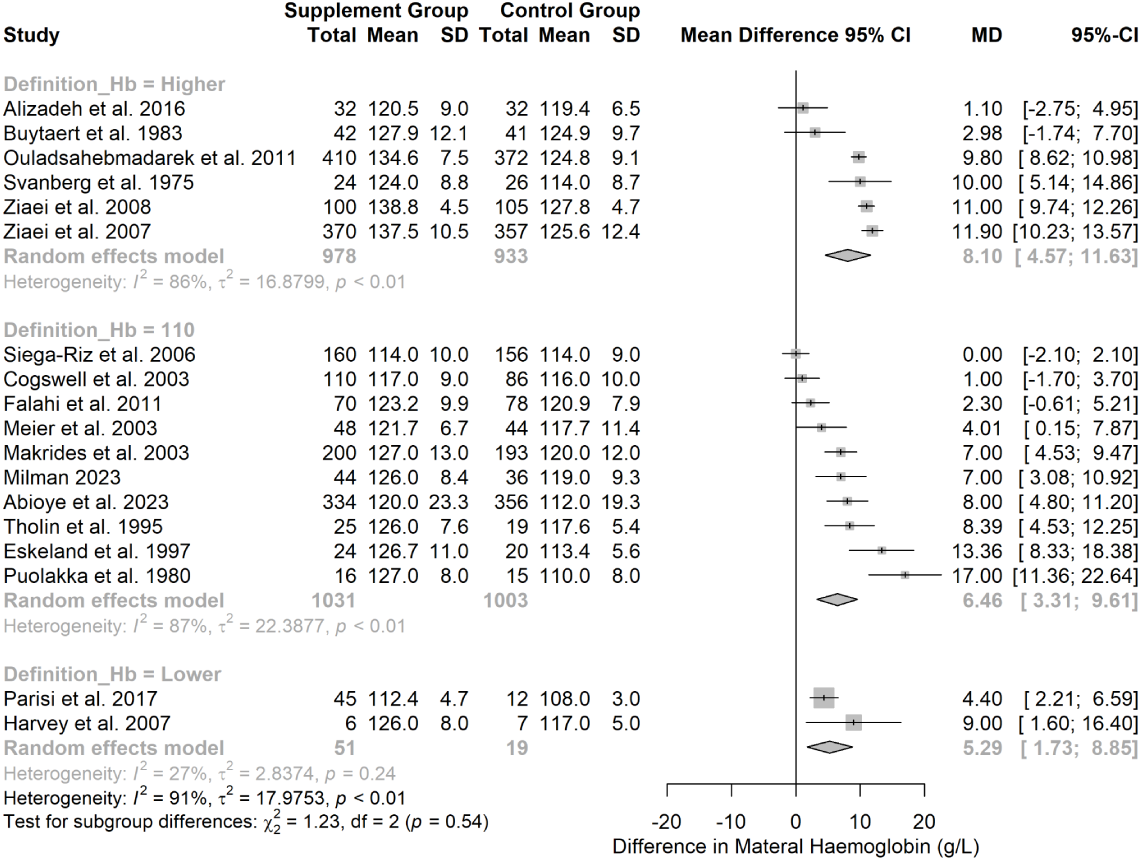
Forest plot showing effect of iron supplementation on maternal haemoglobin with subgroup analysis based on authors’ definition of anaemia (Hb<110g/L, higher or lower haemoglobin value cut offs).

**Figure S5.**
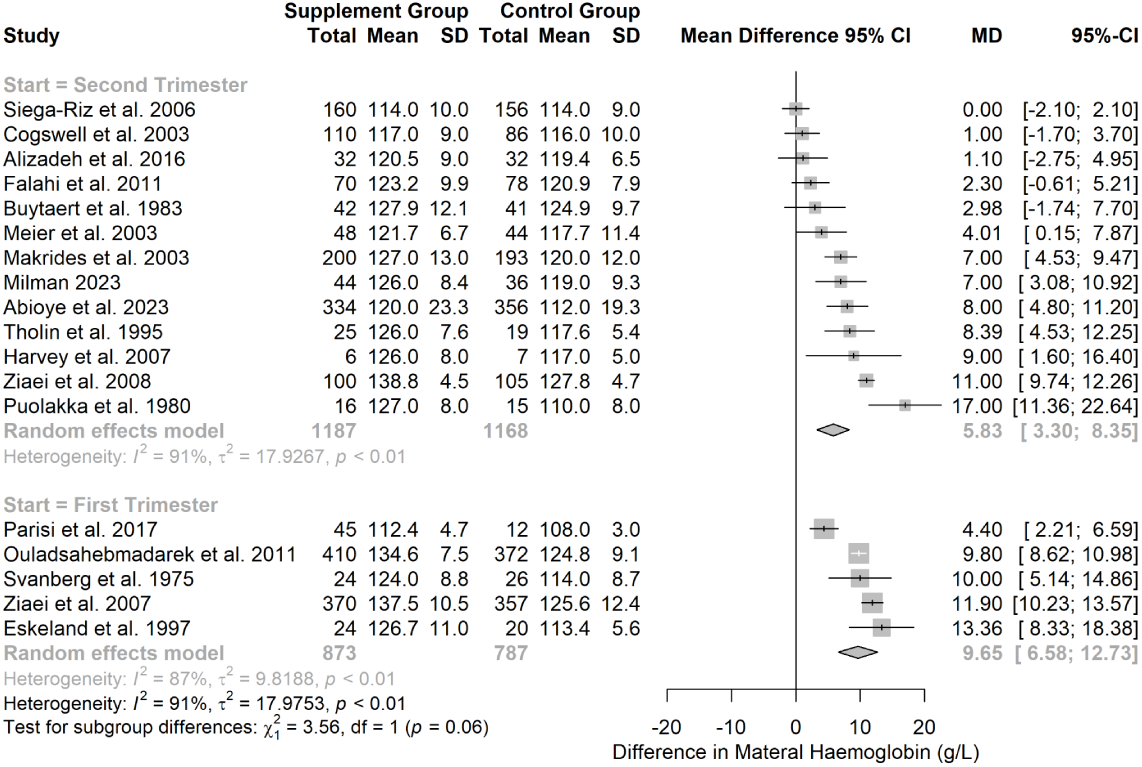
Forest plot showing effect of iron supplementation on maternal haemoglobin with subgroup analysis based on supplement start time (first or second trimester).

**Figure S6.**
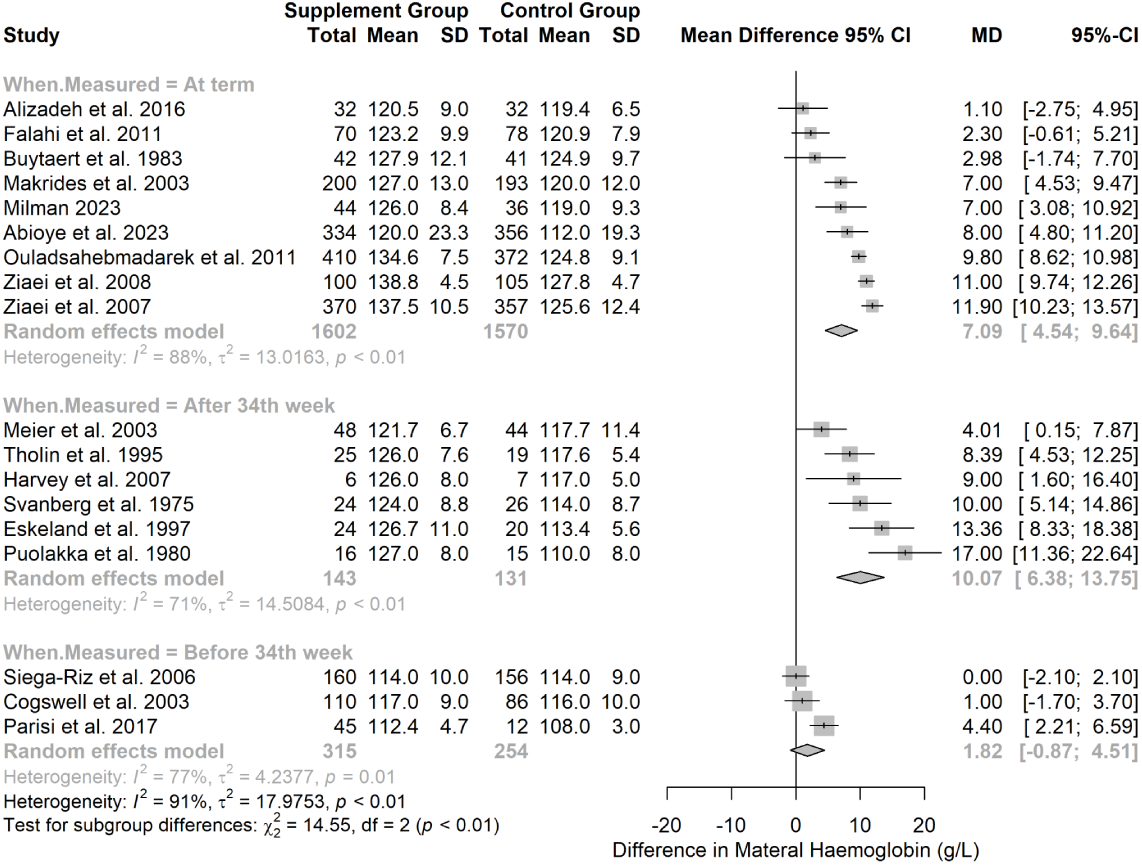
Forest plot showing effect of iron supplementation on maternal haemoglobin with subgroup analysis based on time point of outcome measurement (<34 weeks, 34 to 40 weeks or at term). Significant subgroup differences were detected between studies measuring haemoglobin before and after 34 weeks (Chi^2^ = 14.55, p < .01).

**Figure S7.**
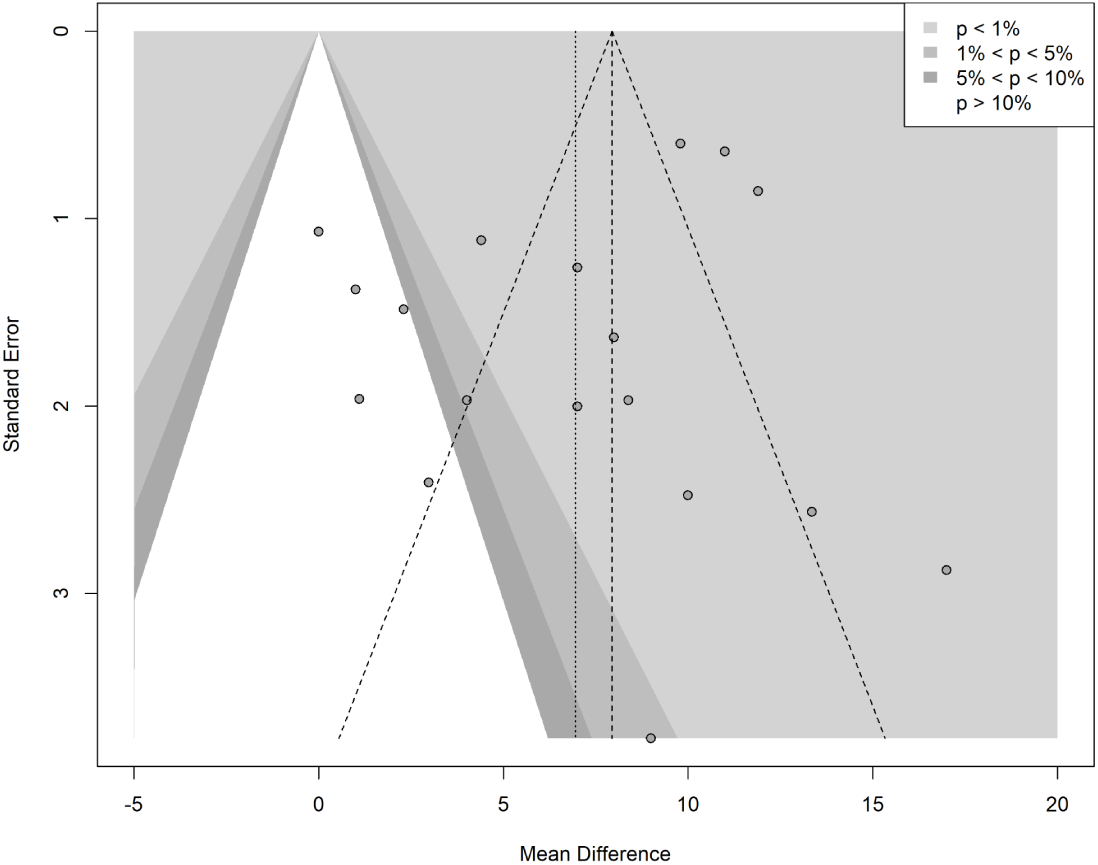
Funnel plot for the effect of iron supplementation on maternal haemoglobin with p-value contours.

**Figure S8.**
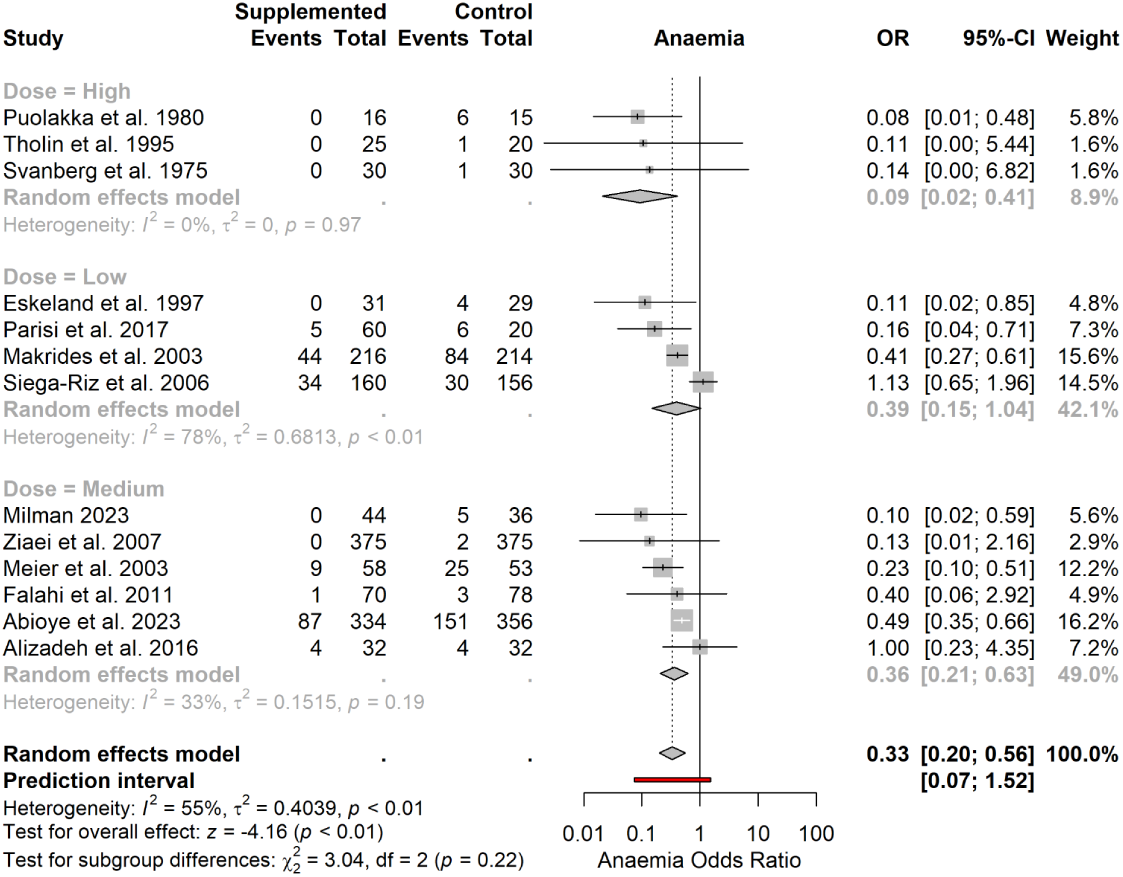
Forest plot showing effect of iron supplementation on odds of maternal anaemia with subgroup analysis based on supplement dose (<31mg, 31mg to 60mg, >60mg).

**Figure S9.**
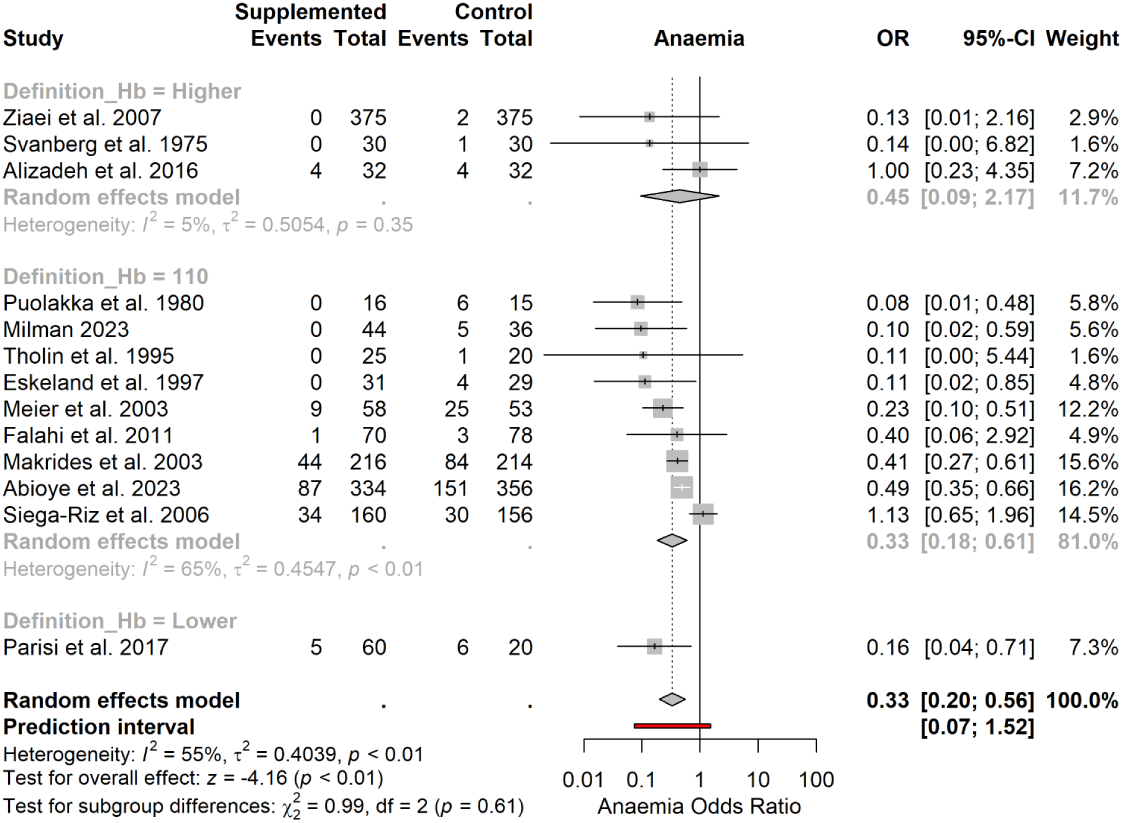
Forest plot showing effect of iron supplementation on odds of maternal anaemia with subgroup analysis based on authors’ definition of anaemia (Hb<110g/L, higher or lower haemoglobin value cut offs).

**Figure S10.**
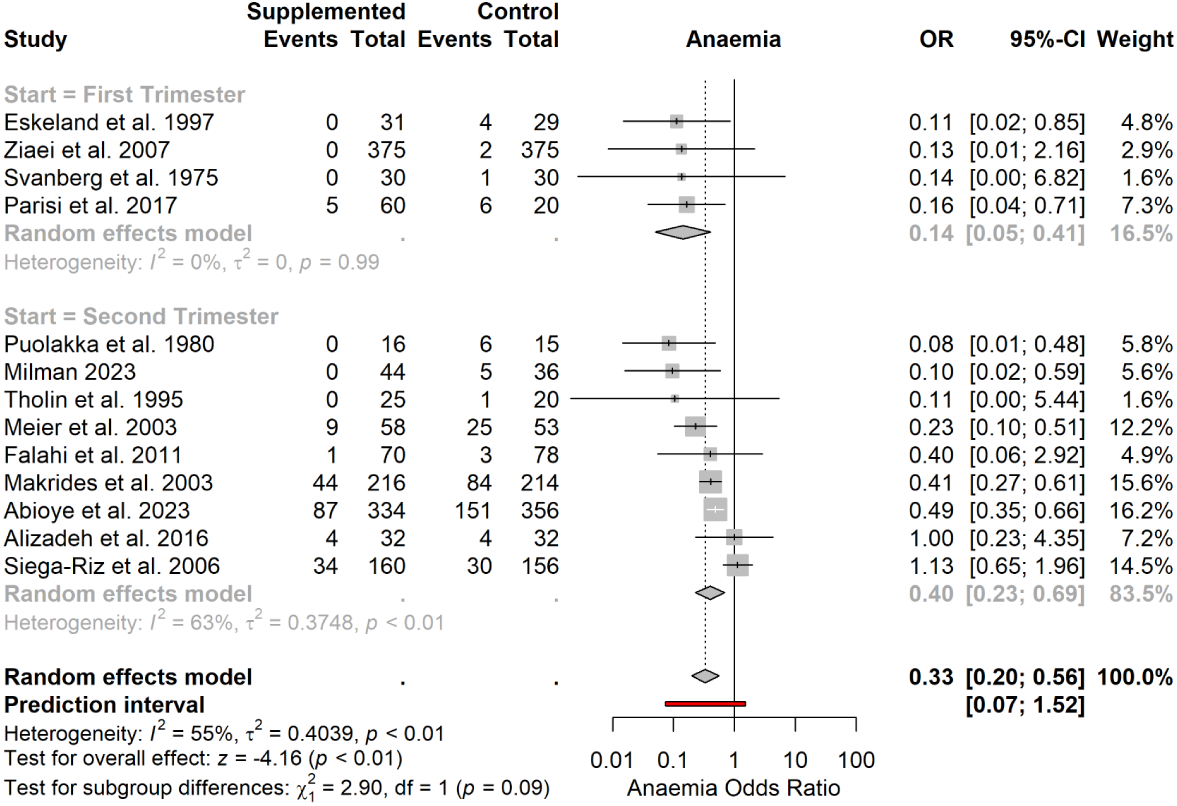
Forest plot showing effect of iron supplementation on odds of maternal anaemia with subgroup analysis based on supplement start time (first or second trimester).

**Figure S11.**
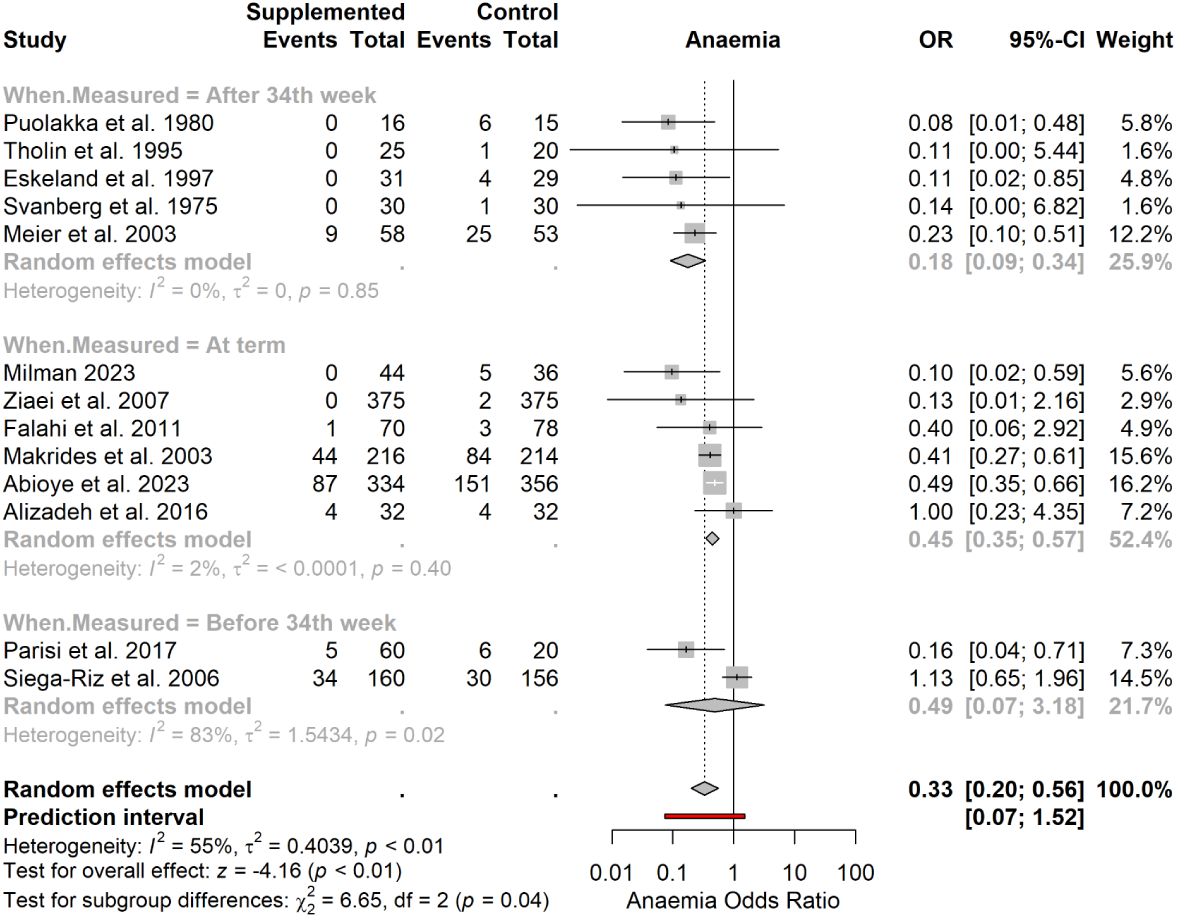
*Forest plot showing effect of iron supplementation on odds of maternal anaemia with subgroup analysis based on time point of outcome measurement (<34 weeks, 34 to 40 weeks or at term). Significant subgroup differences were detected between studies* (Chi^2^ = 6.65, p < .04).

**Figure S12.**
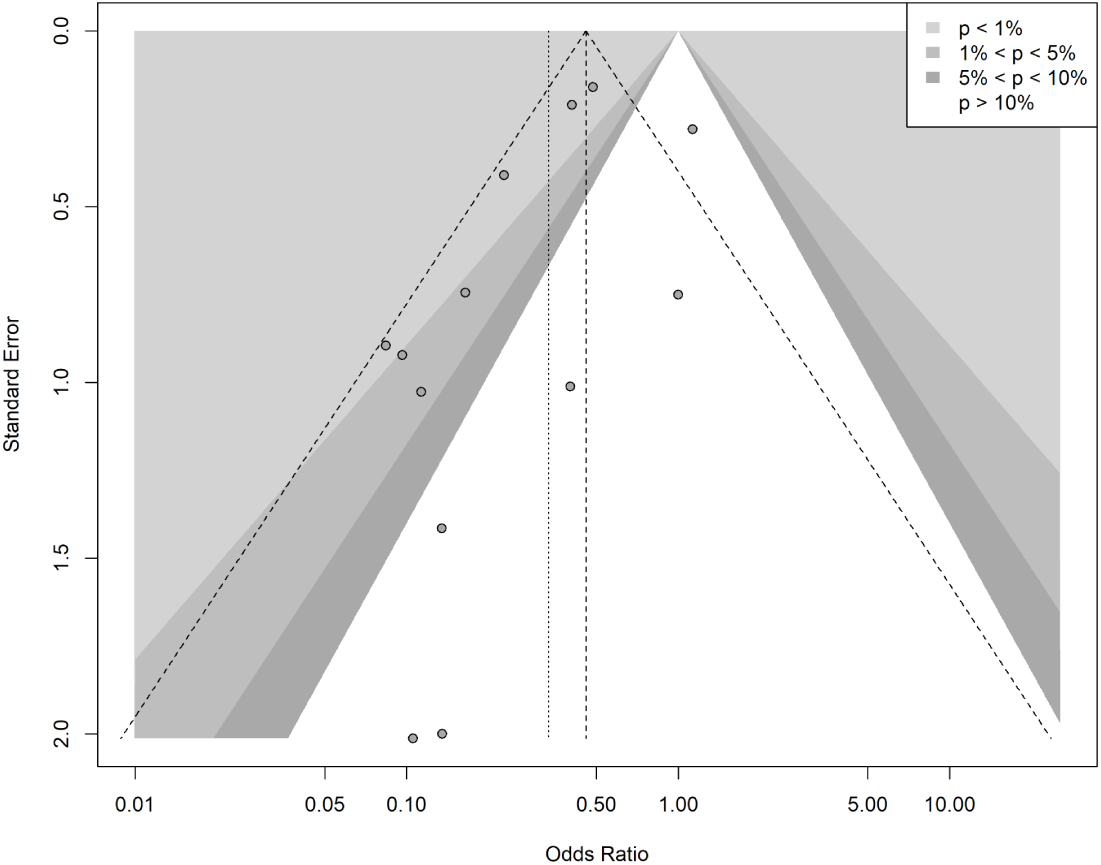
Funnel plot for the effect of iron supplementation on odds of maternal anaemia with p-value contours.

**Figure S13.**
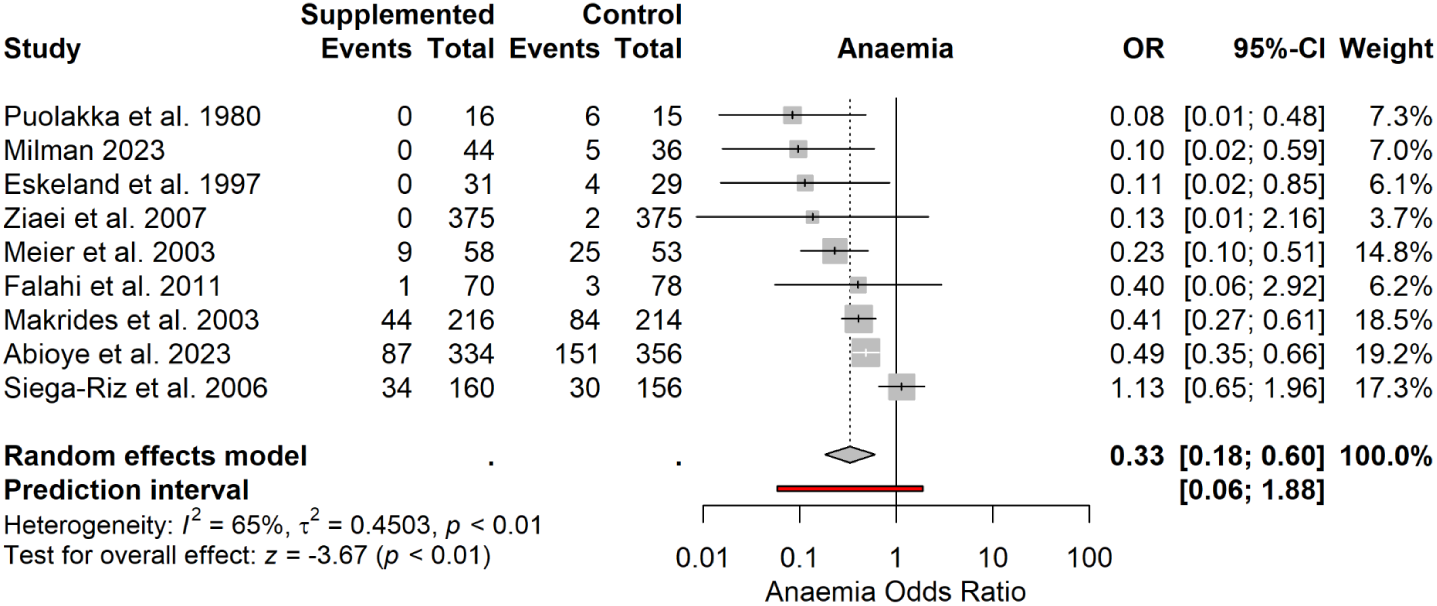
Forest plot showing effect of iron supplementation on odds of maternal anaemia with all “high risk of bias” studies removed.

**Figure S14.**
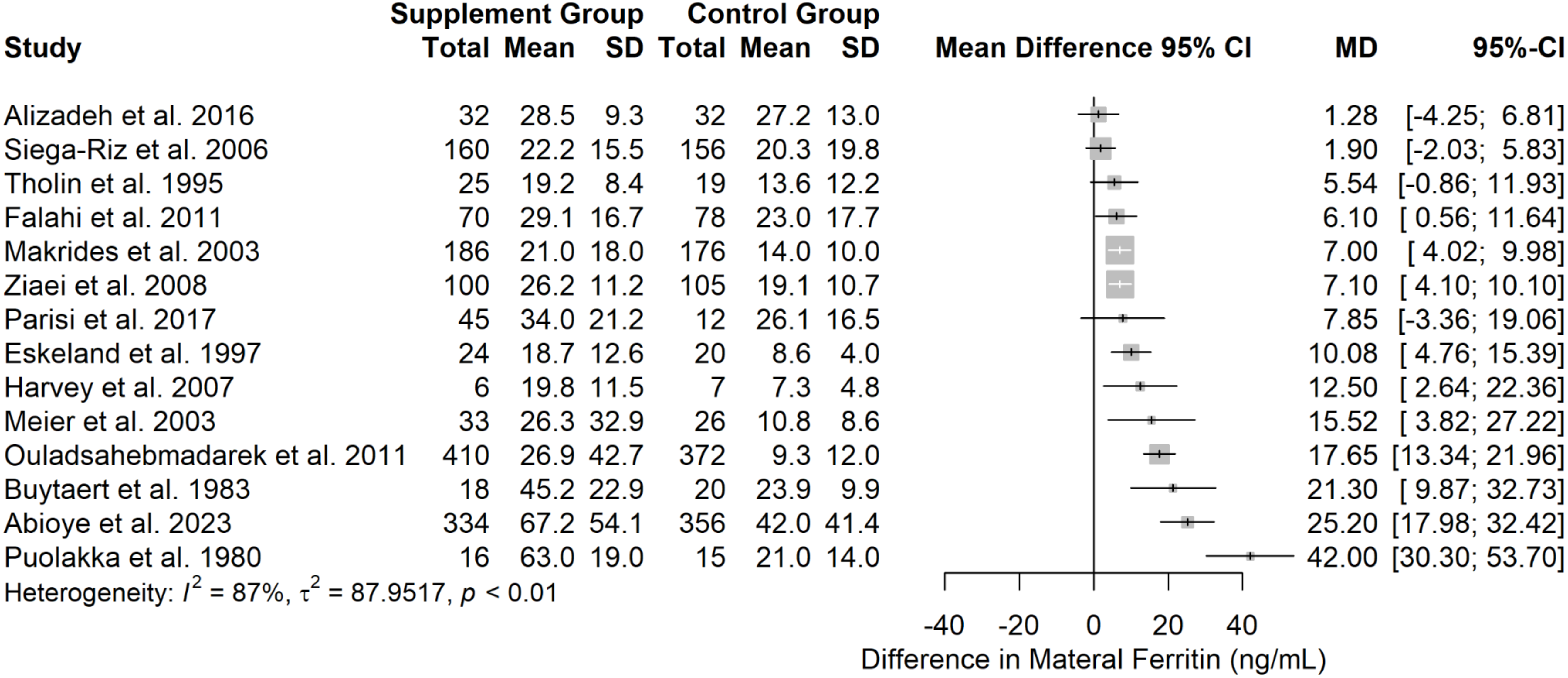
Forest plot showing effect of iron supplementation on maternal ferritin. A pooled effect estimate is not reported due to the high degree of heterogeneity (I^2^ = 87%).

**Figure S15.**
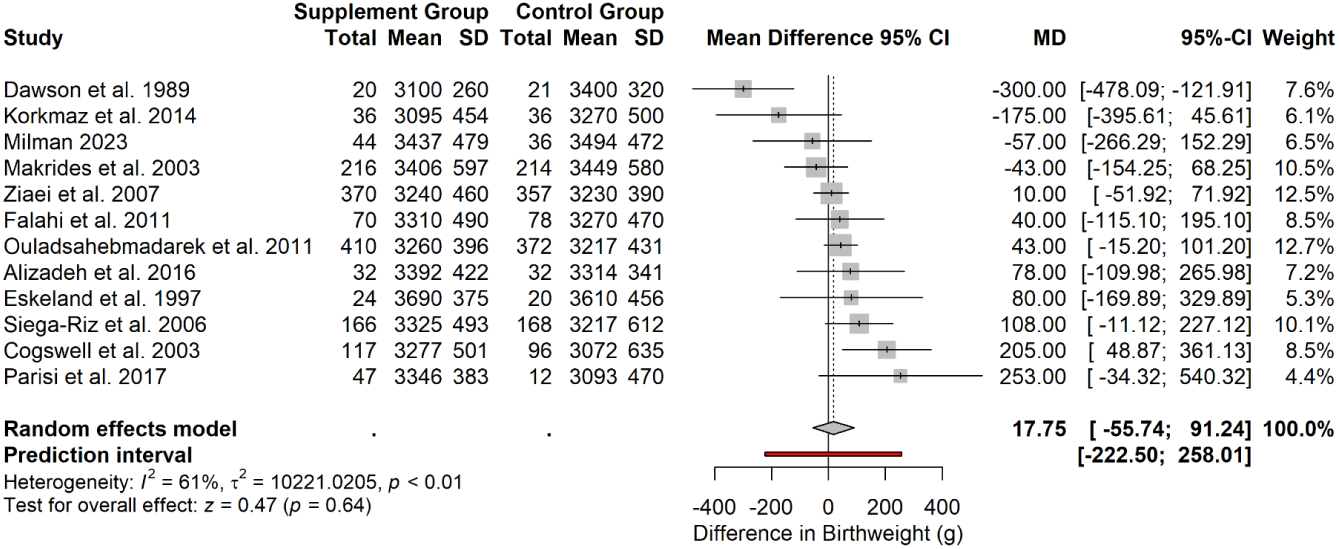
Forest plot showing effect of iron supplementation on foetal birth weight.

**Figure S16.**
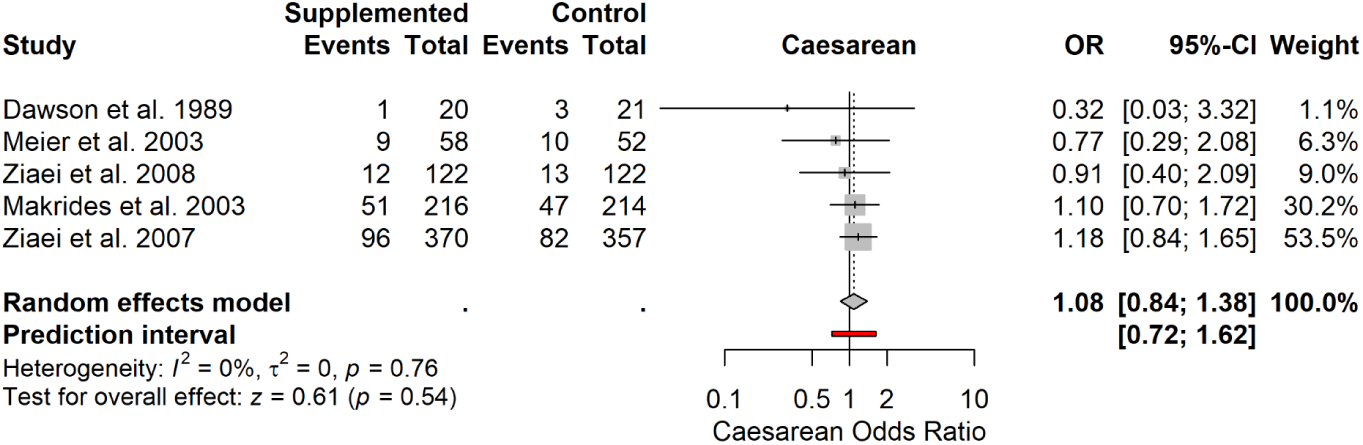
Forest plot showing effect of iron supplementation on odds of caesarean section.

**Figure S17.**
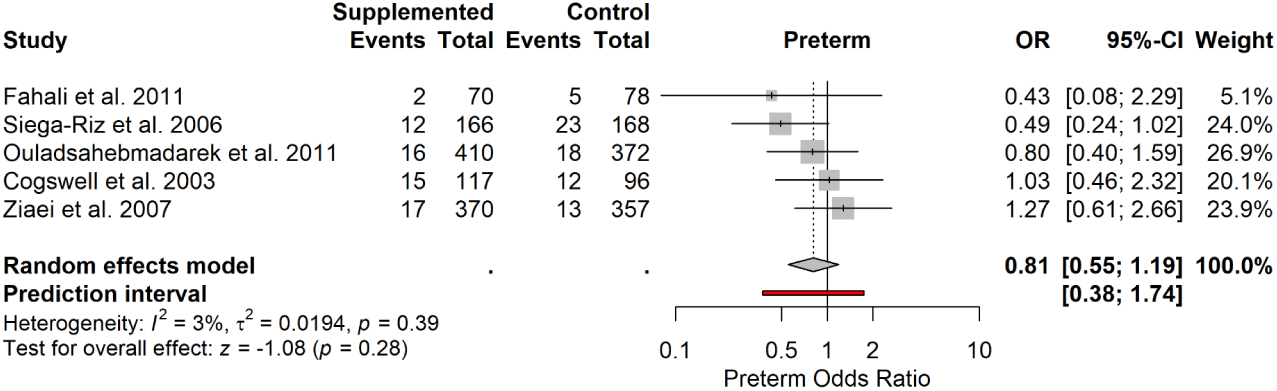
Forest plot showing effect of iron supplementation on odds of preterm birth.

